# COVID-19 severity in asthma patients: A multi-center matched cohort study

**DOI:** 10.1101/2020.10.02.20205724

**Authors:** Lacey B. Robinson, Liqin Wang, Xiaoqing Fu, Zachary S. Wallace, Aidan A. Long, Yuqing Zhang, Carlos A. Camargo, Kimberly G. Blumenthal

## Abstract

**Objective:** The evidence pertaining to the effects of asthma on Coronavirus disease 2019 outcomes has been unclear. To improve our understanding of the clinically important association of asthma and Coronavirus disease 2019.

**Methods:** A matched cohort study was performed using data from the Mass General Brigham Health Care System (Boston, MA). Adult (age ≥ 18 years) patients with confirmed Coronavirus disease 2019 and without chronic obstructive pulmonary disease, cystic fibrosis, or interstitial lung disease between March 4, 2020 and July 2, 2020 were analyzed. Up to 5 non-asthma comparators were matched to each asthma patient based on age (within 5 years), sex, and date of positive test (within 7 days). The primary outcomes were hospitalization, mechanical ventilation, and death, using multivariable Cox-proportional hazards models accounting for competing risk of death, when appropriate. Patients were followed for these outcomes from diagnosis of Coronavirus disease 2019 until July 2, 2020.

**Results:** Among 562 asthma patients, 199 (21%) were hospitalized, 15 (3%) received mechanical ventilation, and 7 (1%) died. Among the 2686 matched comparators, 487 (18%) were hospitalized, 107 (4%) received mechanical ventilation, and 69 (3%) died. The adjusted Hazard Ratios among asthma patients were 0.99 (95% Confidence Internal 0.80, 1.22) for hospitalization, 0.69 (95% Confidence Internal 0.36, 1.29) for mechanical ventilation, and 0.30 (95% Confidence Internal 0.11, 0.80) for death.

**Conclusions:** In this matched cohort study from a large Boston-based healthcare system, asthma was associated with comparable risk of hospitalization and mechanical ventilation but a lower risk of mortality.

## INTRODUCTION

Coronavirus disease 2019 (COVID-19), is caused by the novel severe acute respiratory syndrome coronavirus 2 (SARS-CoV-2), and is an ongoing continued global health crisis.^1^ The United States (US) became an epicenter for COVID-19 and Boston, Massachusetts experienced a significant epidemic in Spring 2020.^2^

Viral respiratory illnesses are known to be associated with asthma exacerbations and severe outcomes,^3^ but relatively little is known about the severity of COVID-19 among asthma patients. Prior, smaller outbreaks of novel coronavirus, including Severe Acute Respiratory Syndrome (SARS) in 2002 and Middle Eastern Respiratory Syndrome (MERS) in 2012 -- had similar severe clinical outcomes to COVID-19, but severe disease among asthma patients was variable.^4,5^ While current data from the COVID-19 pandemic is limited, there have been conflicting reports on the association of asthma to severity of COVID-19.^6-8^

Whether asthma is associated with severe clinical outcomes in COVID-19 is of high clinical importance to the over 20 million adult patients with asthma in the US.^9^ We therefore performed a matched cohort study to understand the relation of asthma to COVID-19 disease severity in patients with SARS-CoV-2 infection in a large Boston-based health care system.

## METHODS

### Study Population

Mass General Brigham (MGB, formerly Partners HealthCare System) is a large health care system that includes tertiary care hospitals (Massachusetts General Hospital and Brigham and Women’s Hospital), community hospitals, and primary and specialty outpatient centers in the greater Boston area. We used data from three sources from MGB: the MGB Enterprise Data Warehouse, MGB Research Patient Data Registry, and the COVID-19 Datamart.^10^ All data sources were linked by the Enterprise Master Patient Index, a unique patient identifier.

We identified patients seen at MGB who were ≥ 18 years of age and had a positive test result for SARS-CoV-2 by polymerase chain reaction (PCR) clinical assay between March 4, 2020 and July 2, 2020. From these patients, we excluded patients with non-asthma chronic lung diseases including chronic obstructive pulmonary disease (COPD), cystic fibrosis (CF), and interstitial lung disease (ILD) by *International Classification of Diseases, Tenth Revision, Clinical Modification* (ICD-10-CM) code (**eTable 1**).

### Exposure: Asthma

We defined asthma based on the following criteria: 1) A patient with ≥ 2 diagnosis codes for asthma by ICD-10-CM (**eTable 1**) and 2) prescription of an asthma medication (including short-acting beta agonist, long-acting beta agonist, inhaled-corticosteroid, and montelukast) in the one year prior to diagnosis of COVID-19. The asthma case definition was informed by manual review by a board-certified allergist/immunologist (L.B.R.); the chosen asthma definition confirmed asthma in 24 of 25 cases (96%) reviewed.

We defined allergic asthma patients as having at least one of the following: 1) history of allergic rhinitis by ICD-10-CM diagnosis code in the last one year (see **eTable 1**) or 2) on therapy with oral antihistamine, leukotriene modifier, intranasal corticosteroid spray, or intranasal antihistamine in the last one year (see **eTable 2**). All asthma patients who did not meet the definition of allergic asthma were classified as non-allergic asthma.

We defined severe asthma as: 1) used asthma biologics (anti-IgE, anti-interleukin-5/interleukin-5 receptor, or anti-interleukin-4 receptor) in the last one year, or 2) received oral corticosteroids ≥3 times in the last one year, or 3) received theophylline in the last one year. All asthma patients who did not meet the definition of severe asthma were classified as non-severe asthma.

### Comparators

For each asthma patient, we identified up to five SARS-CoV-2 infected comparator patients without asthma matched on age group (within 5 years), sex, and date of positive SARS-CoV-2 test (within 7 days). The first positive SARS-CoV-2 test date was used for matching.

### Outcomes: Hospitalization, Mechanical Ventilation, Death

The primary outcomes were hospitalization, mechanical ventilation, and death. Patients were followed for these outcomes from diagnosis of SARS-CoV-2 until July 2, 2020.

### Covariates

We considered multiple covariates including demographics (age, sex, race/ethnicity), expected payor, smoking status, body mass index, and comorbid conditions prior to diagnosis of SARS-CoV-2. We also calculated Charlson Comorbidity Index for each patient (**eTable 3**).^11,12^

### Statistical Analyses

Categorical variables are presented as number (percentage), and continuous variables are reported as mean ± standard deviation or median ± interquartile range, as appropriate. Continuous variables were compared using a two-sample t-test for continuous normally distributed variables or Mann-Whitney U Test for continuous non-normally distributed variables. Categorical variables were compared using Chi-square tests or Fisher’s exact test, as appropriate.

Person-days of follow-up for each patient were computed as the amount of time from the date of first positive SARS-CoV-2 test to the date of the hospitalization (or mechanical ventilation, or death) or until July 2, 2020. We examined the relation of asthma to COVID-19 outcomes using multivariable-adjusted Cox-proportional hazards models; for the outcomes of hospitalization and mechanical ventilation, models accounted for competing risk of death. Multivariable models included covariates selected *a priori*. We took the same approach to examine relation of allergic vs. non-allergic asthma as well as the severe vs. non-severe asthma to the risk of COVID-19 outcomes.

We also conducted a sensitivity analysis to assess for the effect of smoking status by excluding smokers. Statistical analyses were completed using SAS (version 9.4; SAS Institute, Inc.) with a two-tailed *P* < 0.05 considered statistically significant.

### Institutional Review Board

This study was reviewed by the Partners Human Research Committee and determined to be exempt/non-human subjects research (Protocol 2020P000833).

## RESULTS

We identified 562 patients with asthma and 2686 non-asthma comparators matched on age, sex and date of positive SARS-CoV-2. Asthma patients differed from matched comparators on several characteristics including race, ethnicity, and expected payor (**Table 1**). Mean body mass index was higher among asthma patients than comparators (31.8 kg/m^2^ vs. 29.8 kg/m^2^, respectively). Asthma patients had more comorbid conditions including diabetes and hypertension.

**Table 1.**
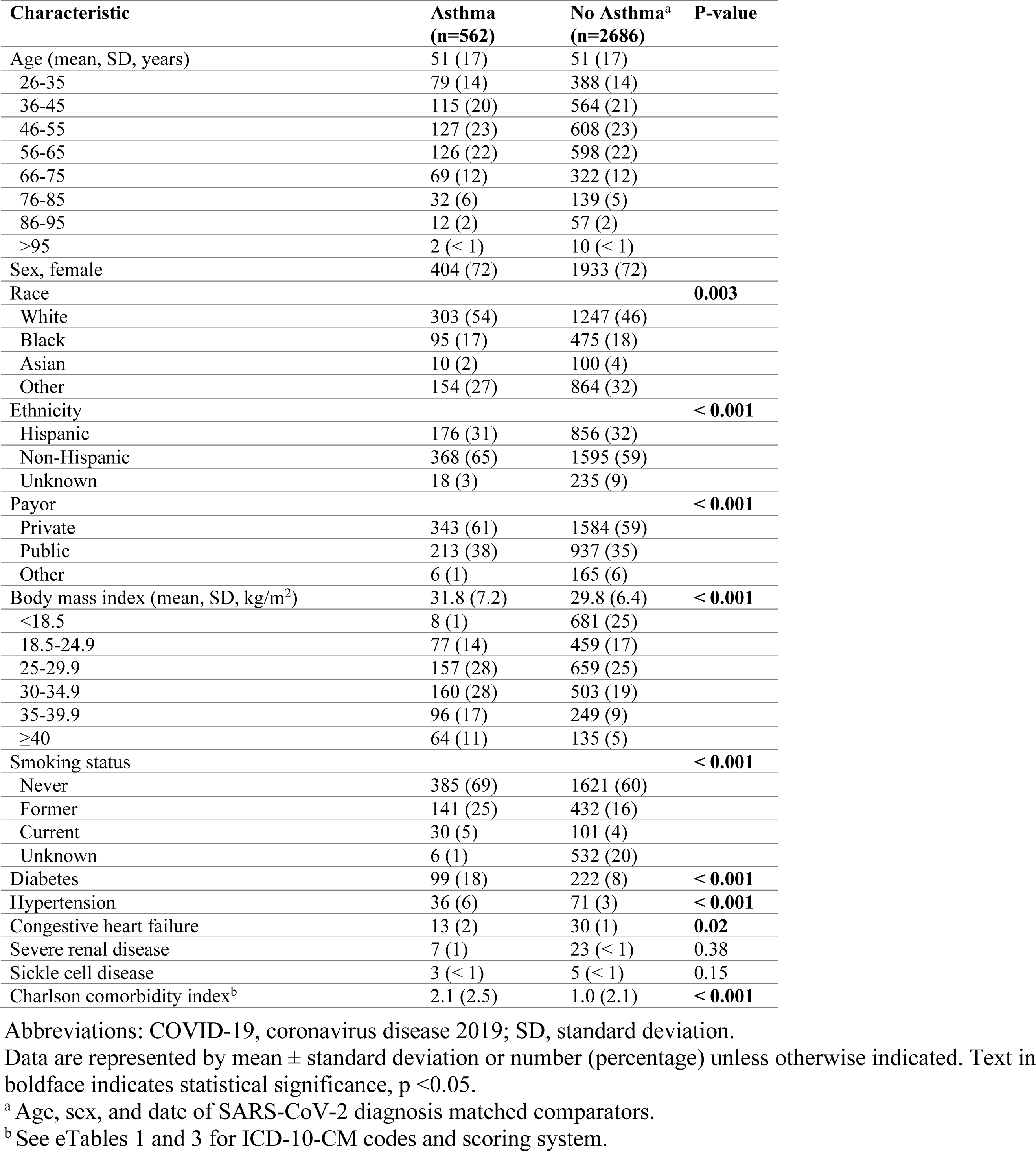
Clinical characteristics of COVID-19 patients with and without asthma

Hospitalization occurred in 119 (21%, 3.7 per 1000 person days) asthma patients and 487 (18%, 3.0 per 1000 person days) matched comparators (**Table 2**). In the fully adjusted model, the rate of hospitalization did not differ significantly between the two groups (adjusted hazard ratio [aHR] 0.99, 95%CI: 0.80, 1.22). Mechanical ventilation was utilized in 15 (3%, 0.4 per 1000 person days) asthma patients and 107 (4%, 0.6 per 1000 person days) comparators. In the fully adjusted model, the rate of mechanical ventilation was not significantly different between the two groups (aHR 0.69, 95%CI: 0.36,1.29). Death occurred in 7 (1%, 0.2 per 1000 person days) asthma patients and 69 (3%, 0.3 per 1000 person days) comparators. In the fully adjusted model, asthma was associated with a lower risk of death (aHR 0.30, 95%CI: 0.11, 0.80).

**Table 2.** Clinical outcomes of COVID-19 patients with asthma and without asthma

| **Outcomes** | **Asthma (n=562)** | **No Asthma (n=2686)<sup>a</sup>** |
| --- | --- | --- |
| **Hospitalization:** |  |  |
| No. of events | 119 (21) | 487 (18) |
| Person days of follow up (Mean, SD) | 58 (34) | 61 (33) |
| Rate of event per 1000 person days (95%CI) | 3.7 (3.0, 4.3) | 3.0 (2.7, 3.3) |
| Matched unadjusted (95%CI)<sup>a</sup> | 1.15 (0.96, 1.38) | 1.00 (ref) |
| Partially adjusted hazard ratio (95%CI)<sup>b</sup> | 1.07 (0.88, 1.29) | 1.00 (ref) |
| Fully adjusted hazard ratio (95%CI)<sup>c</sup> | 0.99 (0.80, 1.22) | 1.00 (ref) |
| **Mechanical Ventilation:** |  |  |
| No. of events | 15 (3) | 107 (4) |
| Person days of follow up (Mean, SD) | 71 (23) | 71 (24) |
| Rate of event per 1000 person days (95%CI) | 0.4 (0.2, 0.6) | 0.6 (0.5, 0.7) |
| Matched unadjusted (95%CI)<sup>a</sup> | 0.67 (0.39, 1.15) | 1.00 (ref) |
| Partially adjusted hazard ratio (95%CI)<sup>b</sup> | 0.65 (0.37, 1.15) | 1.00 (ref) |
| Fully adjusted hazard ratio (95%CI)<sup>c</sup> | 0.69 (0.36, 1.29) | 1.00 (ref) |
| **Death:** |  |  |
| No. of events | 7 (1) | 69 (3) |
| Person days of follow up (Mean, SD) | 73 (21) | 73 (21) |
| Rate of event per 1000 person days (95%CI) | 0.2 (0.0, 0.3) | 0.3 (0.3, 0.4) |
| Matched unadjusted (95%CI)<sup>a</sup> | 0.47 (0.22, 1.02) | 1.00 (ref) |
| Partially adjusted hazard ratio (95%CI)<sup>b</sup> | 0.50 (0.22, 1.13) | 1.00 (ref) |
| Fully adjusted hazard ratio (95%CI)<sup>c</sup> | **0.30 (0.11, 0.80)** | 1.00 (ref) |

Abbreviations: COVID-19, coronavirus disease 2019; SD, standard deviation; CI, confidence interval.
Text in boldface indicates statistical significance, $p < 0.05$ .
<sup>a</sup> Age, sex and, date of SARS-CoV-2 diagnosis matched comparators.
<sup>b</sup> Matched on age, sex, and date of SARS-CoV-2 test date and adjusted for age, sex, race, ethnicity, payor, and smoking status.
<sup>c</sup> Matched on age, sex, and date of SARS-CoV-2 test date and adjusted for age, sex, race, ethnicity, payor, smoking status, body mass index, and Charlson comorbidity index.

The seven deceased asthma patients ranged in age from 54 to 84 years and 4 (57%) were female. All of these patients had multiple significant co-morbid conditions, and all were treated with inhaled corticosteroids with or without long-acting beta agonist prior to COVID-19 diagnosis.

In a sensitivity analysis excluding all current smokers, among 532 asthma patients and 2542 matched comparators, asthma was not associated with hospitalization or mechanical ventilation, and remained associated with a lower risk of mortality (**eTable 4**).

### Allergic and Non-Allergic Asthma

Among asthma patients, we identified 260 allergic asthma patients and 302 non-allergic asthma comparators. Patient characteristics are described in **eTable 5**. Among allergic asthma patients, we observed a similar rate of hospitalization, mechanical ventilation, and death as compared with their matched comparators (**Table 3**).

**Table 3.**
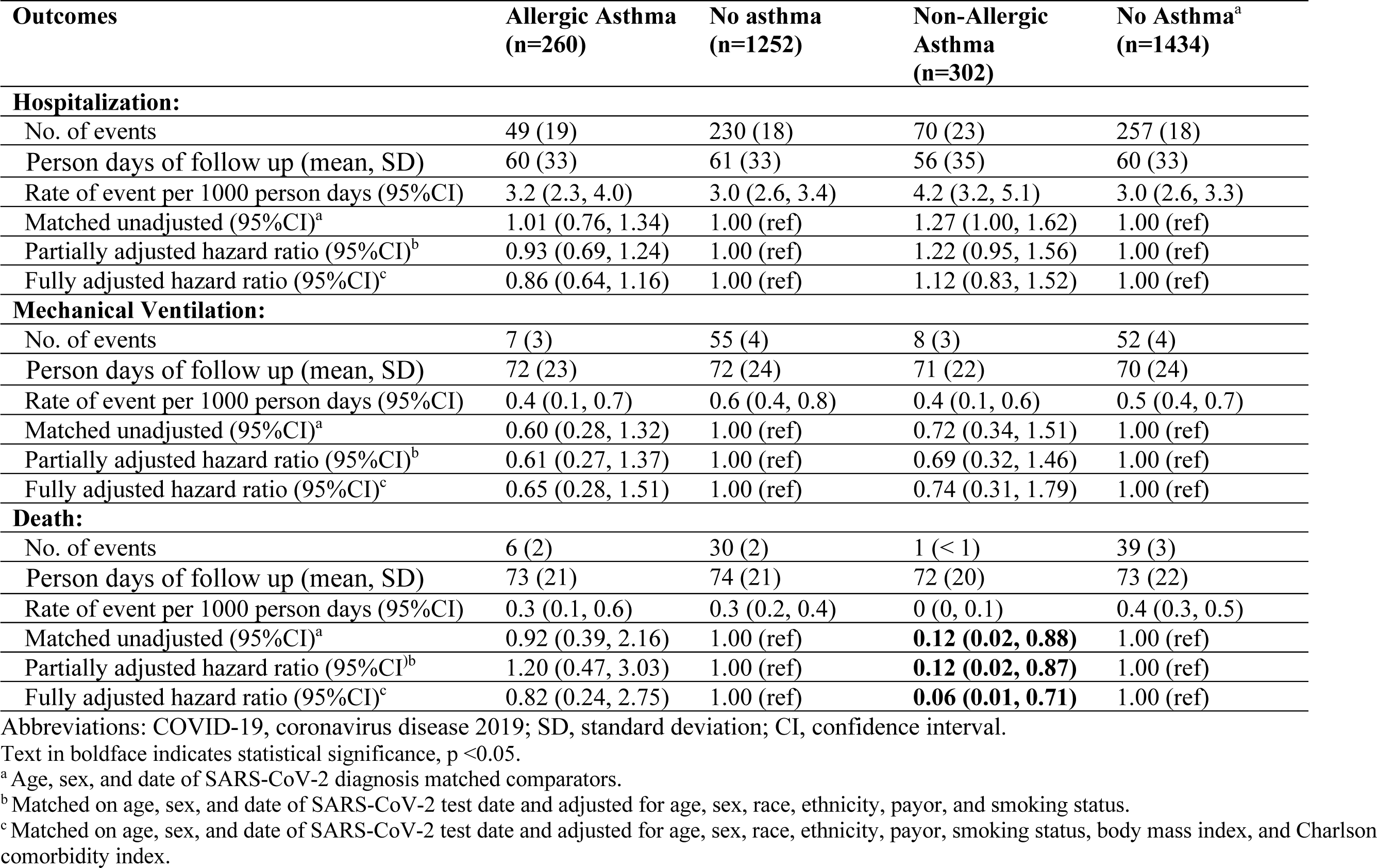
Severe clinical outcomes of COVID-19 patients with allergic and non-allergic asthma and matched comparators

### Severe and Non-severe Asthma

Among asthma patients, we identified 44 patients with severe asthma and 518 comparators with non-severe asthma. Asthma medication use in severe asthma patients included inhaled corticosteroids in 30 patients (68%), inhaled corticosteroids plus long-acting beta agonists in 21 (48%), asthma biologics in 8 (18%), oral corticosteroids ≥ 3 times in the last year in 40 (91%) and theophylline in 1 (2%) (**eTable 6**). In severe asthma patients, hospitalization occurred in 14 (32%, 5.9 per 1000 person days) and mechanical ventilation in 5 (11%, 1.6 per 1000 person days). There were no deaths among severe asthma patients (**Table 4**). There were no significant differences observed in the rates of severe outcomes among severe asthmatics and their matched comparators.

**Table 4.** Severe clinical outcomes of COVID-19 patients with severe and non-severe asthma and age, sex, and diagnosis date matched comparators

| Outcomes | Severe Asthma<br>(n=44) | No Asthma<br>(n=210) | Non-Severe<br>Asthma<br>(n= 518) | No Asthma<sup>a</sup><br>(n=2476) |
| --- | --- | --- | --- | --- |
| **Hospitalization:** |  |  |  |  |
| No. of events | 14 (32) | 45 (21) | 105 (20) | 442 (18) |
| Person days of follow up (mean, SD) | 54 (38) | 61 (36) | 58 (34) | 61 (33) |
| Rate of event per 1000 person days (95%CI) | 5.9 (2.8, 9.0) | 3.5 (2.5, 4.6) | 3.5 (2.8, 4.2) | 2.9 (2.7, 3.2) |
| Matched unadjusted (95%CI)<sup>a</sup> | 1.53 (0.89, 2.64) | 1.00 (ref) | 1.12 (0.92, 1.36) | 1.00 (ref) |
| Partially adjusted hazard ratio (95%CI)<sup>b</sup> | 1.73 (0.98, 3.04) | 1.00 (ref) | 1.03 (0.84, 1.26) | 1.00 (ref) |
| Fully adjusted hazard ratio (95%CI)<sup>c</sup> | 1.99 (0.82, 4.79) | 1.00 (ref) | 0.94 (0.75, 1.17) | 1.00 (ref) |
| **Mechanical Ventilation:** |  |  |  |  |
| No. of events | 5 (11) | 12 (6) | 10 (2) | 95 (4) |
| Person days of follow up (mean, SD) | 71 (26) | 73 (25) | 71 (22) | 71 (24) |
| Rate of event per 1000 person days (95%CI) | 1.6 (0.2, 3.0) | 0.8 (0.3, 1.2) | 0.3 (0.1, 0.4) | 0.5 (0.4, 0.7) |
| Matched unadjusted (95%CI)<sup>a</sup> | 1.95 (0.75, 5.11) | 1.00 (ref) | **0.49 (0.26, 0.95)** | 1.00 (ref) |
| Partially adjusted hazard ratio (95%CI)<sup>b</sup> | 2.10 (0.77, 5.76) | 1.00 (ref) | **0.47 (0.24, 0.92)** | 1.00 (ref) |
| Fully adjusted hazard ratio (95%CI)<sup>c</sup> | 85.2 (5.55, 1310) | 1.00 (ref) | 0.47 (0.22, 1.01) | 1.00 (ref) |
| **Death:** |  |  |  |  |
| No. of events | 0 (0) | 9 (4) | 7 (1) | 60 (2) |
| Person days of follow up (mean, SD) | 78 (17) | 76 (21) | 72 (21) | 73 (21) |
| Rate of event per 1000 person days (95%CI) | - | 0.6 (0.2, 0.9) | 0.2 (0, 0.3) | 0.3 (0.2, 0.4) |
| Matched unadjusted (95%CI)<sup>a</sup> | - | 1.00 (ref) | 0.55 (0.25, 1.20) | 1.00 (ref) |
| Partially adjusted hazard ratio (95%CI)<sup>b</sup> | - | 1.00 (ref) | 0.58 (0.26, 1.33) | 1.00 (ref) |
| Fully adjusted hazard ratio (95%CI)<sup>c</sup> | - | 1.00 (ref) | **0.34 (0.12, 0.96)** | 1.00 (ref) |

Abbreviations: COVID-19, coronavirus disease 2019; SD, standard deviation; CI, confidence interval.
Text in boldface indicates statistical significance, $p < 0.05$ .
<sup>a</sup> Age, sex, and date of SARS-CoV-2 diagnosis matched comparators and adjusted for age, sex, race, ethnicity, payor, and smoking status.
<sup>b</sup> Matched on age, sex, and date of SARS-CoV-2 test date.
<sup>c</sup> Matched on age, sex, and date of SARS-CoV-2 test date and adjusted for age, sex, race, ethnicity, payor, smoking status, body mass index, and Charlson comorbidity index.

## DISCUSSION

In this multi-center matched cohort study, we found that asthma patients with COVID-19 were at similar risk of hospitalization and mechanical ventilation as comparators after matching on age, sex and date of SARS-CoV-2 positive test and multivariable adjustment. By contrast, and despite consideration of asthma as potential high-risk comorbid disease, asthma patients were at a 3-fold significantly lower risk of mortality compared to their non-asthma comparators, a finding that persisted even in a sensitivity analysis that excluded active smokers. We did not observe any clear risk of severe COVID-19 outcomes when we considered clinical outcomes by asthma phenotype (allergic and non-allergic asthma) or severity (severe and non-severe asthma).

Viral respiratory illnesses are known triggers of asthma exacerbations and asthma patients are at increased risk of severe respiratory infections.^3^ Severe disease among asthma patients in prior novel coronavirus outbreaks was variable. In the SARS outbreak, asthma was over-represented in cases and associated with severe disease.^4^ Conversely, in the MERS outbreak, asthma was not overrepresented in cases and not associated with severe disease.^5^ The data from prior outbreaks is limited due to the smaller number of infected patients and relatively short duration of outbreak.

COVID-19 has spurred a pandemic of illness that is unprecedented in recent times. The relationship between asthma and severe outcomes has not been well defined. Initial reports from Wuhan, China, suggested that asthma was not a risk factor for severe outcomes in COVID-19 ^13,14^ and additional early reports suggested that asthma was unrepresented among severe cases.^15^ However, initial reports from New York City found a high prevalence of asthma among COVID-19 patients.^7^ This, coupled with our prior knowledge of severe viral illnesses in asthma patients, prompted the Centers for Disease Control and Prevention to consider asthma a potential risk factor for severe COVID-19.^16^ However, subsequent reports from around the world have suggested that asthma may not be risk factor for severe COVID-19, including hospitalization and death.^6,8,17-19^ However, these reports have largely been limited by several factors including the inclusion of other chronic pulmonary conditions (e.g. COPD) in populations studied, conditioning on patients who were hospitalized with COVID-19,^18^ and/or not accounting for secular changes in COVID-19 testing criteria.^20^ Our study improves upon these prior studies by rigorously defining asthma, excluding those with non-asthma chronic pulmonary conditions (i.e. COPD, CF, and ILD) who appear to be higher risk of severe COVID-19 outcomes with the risk being primarily driven by COPD,^21,22^ and matching by PCR testing date. Together, this facilitates a more precise assessment of the true relation between asthma and severe outcomes in COVID-19.

Our data indicates that asthma patients with COVID-19 are equally likely to be hospitalized and receive mechanical ventilation but that death was significantly less likely. Although the factors underlying these findings are not yet known, important considerations include: possible biologic mechanisms (e.g. angiotensin-converting enzyme 2 [ACE2] receptor expression and TH2 cytokines) and possible protective effects of asthma medications (e.g. corticosteroids).

With regard to the ACE2 receptor, the novel SARS-CoV-2 virus enters target cells by binding to this ACE2 receptor in the nasal mucosa and type II pneumocytes in the lungs.^23^ ACE2 receptor expression is variable and can be upregulated by anti-viral interferons.^23^ There appears to be additional variability among allergic asthma patients and in the presence of Th2 cytokines such as interleukin-13.^24,25^ Indeed, a recent study performed using population-based data from the United Kingdom Biobank reported no significantly elevated risk of severe COVID among allergic asthmatics.^22^ In our study, we report a similar risk of severe COVID-19 outcomes among allergic asthma patients and their comparators. However, it is possible that we were unable to define all allergic asthma patients due to limitations of case identification by ICD-10-CM codes and anti-allergic medications, many of which are over-the-counter pharmaceuticals.

The primary treatment for patients with persistent asthma is inhaled corticosteroids. There is limited evidence regarding the relation between inhaled corticosteroids and COVID-19; however, there is some evidence to suggest therapy may be beneficial.^15,26^ For example, ciclesonide, an inhaled corticosteroid, has the potential to inhibit viral replication of SARS-CoV-2.^27^ Moreover, the only therapy with preliminary evidence to support a reduction in mortality in severe COVID-19 is a corticosteroid – dexamethasone.^28^ A recent rapid meta-analysis attempted to review data on inhaled corticosteroid therapy in SARS, MERS or COVID-19 but found no suitable data for review.^29^ In light of the potential mechanistic benefit and proven benefits of inhaled corticosteroid therapy in asthma patients, the current recommendations encourage continued use of asthma controller therapy (i.e. inhaled corticosteroids) during the COVID-19 pandemic.^16,30^ The effect of inhaled corticosteroids on infection with SARS-CoV-2 and the development of COVID-19 is outside the scope of our current study. However, among our asthma cohort, we observed that most asthma patients were prescribed inhaled corticosteroids prior to diagnosis of COVID-19. Future studies aimed at understanding the relation between asthma therapies and COVID-19 are warranted.

Our study has several limitations, including the use of administrative data which may be affected by coding errors leading to misclassification. However, our use of a strict asthma diagnosis requiring multiple asthma codes in the last one year combined with the requirement of treatment with an asthma medication in the last one year decrease the impact of these potential errors. All patients in our cohort study were diagnosed with COVID-19 by PCR; thus, we are unable to estimate the total effect of asthma on the risk of severe outcomes of COVID-19. Prospective population-based cohort studies including asthmatic and non-asthmatic participants are required to better understand this relationship. Our study may be affected by collider bias because we restricted our study population to only those patients with COVID-19. Finally, confounding by indication bias is of concern due to the fact that testing criteria during the COVID-19 pandemic were at times linked to severity of disease. The effect of this bias was reduced, however, in our study by matching by test date and use of a study period which captured the duration of a COVID-19 epidemic in early 2020 in Boston. Additionally, we did not capture information on treatments for COVID-19 in our study and thus, we are unable to assess the differences related to potential COVID-19 therapeutics. However, all patients were diagnosed within one large hospital system with similar treatment protocols, and there were no approved therapeutics for COVID-19 during the study period’s duration. The use of matching by time of diagnosis additionally limits the likelihood of treatment variability during the study period.

Our study also has several strengths. We used data from a large healthcare system which captures patients from academic and community centers throughout the greater Boston area for the entirety of the first wave of COVID-19 in our area. We have less misclassification of asthma due to definition validation by an allergist and exclusion of other chronic pulmonary conditions (COPD, CF, and ILD). Additionally, by use of matching on date of SARS-CoV-2 test we limit the effects of variable testing criteria on COVID-19 diagnosis. Age is a significant predictor of severe outcomes in COVID-19,^16^ and by matching we are able to better control for age-related confounding.

## CONCLUSION

In summary, we report that among patients diagnosed with SARS-CoV-2 by PCR, asthma patients experienced hospitalization and mechanical ventilation at a similar rate as non-asthmatics but had a lower risk of mortality. In addition, patients with allergic asthma or severe asthma were not at increased risk for severe outcomes compared to their matched comparators. Based on this evidence we suggest that asthma alone should not be considered a risk factor for severe COVID-19.

## Data Availability

The data are from Mass General Brigham and are not publicly available.

## Abbreviations

ACE2: angiotensin-converting enzyme 2
CF: cystic fibrosis
COPD: chronic obstructive pulmonary disease
COVID-19: Coronavirus disease 2019
HR: hazard ratio
ICD-10-CM: International Classification of Diseases, Tenth Revision, Clinical Modification
ILD: interstitial lung disease
MERS: Middle eastern respiratory syndrome
MGB: Mass General Brigham
PCR: polymerase chain reaction
SARS: severe acute respiratory syndrome
SARS-CoV-2: severe acute respiratory syndrome coronavirus 2
US: United States

