## Supplemental Materials for "COVID-19 severity in asthma patients: A multi-center matched cohort study"

**Running title:** COVID-19 and Asthma

**Authors:** Lacey B. Robinson, MD[1,2]; Liqin Wang, PhD[2,3]; Xiaoqing Fu, MS[1,4]; Zachary S. Wallace, MD, MSc[1,2,4]; Aidan A. Long, MD[1,2]; Yuqing Zhang, DSc[1,2,4]; Carlos A. Camargo, Jr, MD DrPH[1,2,4,5]; Kimberly G. Blumenthal, MD MSc[1,2,4]

**Affiliations:**

[1] Division of Rheumatology, Allergy, and Immunology, Massachusetts General Hospital, Boston, MA, USA

[2] Harvard Medical School, Boston, MA, USA

[3] Division of General Internal Medicine and Primary Care, Department of Medicine, Brigham and Women's Hospital, Boston, Massachusetts, USA

[4] Clinical Epidemiology Program, The Mongan Institute, Department of Medicine, Massachusetts General Hospital, Boston, MA, USA

[5]Department of Emergency Medicine, Massachusetts General Hospital, Boston, MA, US

**Supplemental Material Contents:**

- **eTable 1**. Diagnosis codes to identify asthma, allergic asthma, and comorbid conditions
- **eTable 2**. Asthma and allergic rhinitis medications by category
- **eTable 3**. ICD-10-CM codes and scoring system for Charlson comorbidity index
- **eTable 4.** Clinical outcomes of COVID-19 patients with asthma and without asthma: Sensitivity analysis excluding current and former smokers
- **eTable 5.** Characteristics of COVID-19 patients with allergic asthma and non-allergic asthma
- **eTable 6.** Characteristics of COVID-19 patients with severe and non-severe asthma
- **Supplemental References**

### eTable 1. Diagnosis codes to identify asthma, allergic asthma, and comorbid conditions

| Clinical Condition | ICD-10-CM |
|---|---|
| Asthma | J45.x |
| COPD | J41.x, J42, J43.x, or J44.x |
| Cystic Fibrosis | E84.x |
| ILD | J84.x |
| Allergic Rhinitis | J30.1, J30.2, J30.8x, J30.9 |
| Diabetes[1] | E08.x, E09.x, E10.x, E11.x, E13.x |
| Hypertension[2] | I10.x, I11.x, I12.x, I13.x, I15.x |
| Congestive Heart Failure[3] | I11.0, I13.0, I13.2, I25.5, I42.0, I42.5–I42.9, I43, I50.x, P29.0 |
| Severe Renal Disease[3] | I12.0, I13.11, I13.2, N18.5, N18.6, N19, N25.0, Z49.x, Z99.2 |
| Sickle Cell Disease[4] | D57.x |

*Abbreviations*: COPD, chronic obstructive pulmonary disease; ILD, interstitial lung disease.

### eTable 2. Asthma and allergic rhinitis medications by category

| ASTHMA MEDICATIONS | ALLERGIC RHINTIS MEDICATIONS |
|---|---|
| **SABA** | **Antihistamines** |
| Levalbuterol | Fexofenadine |
| Albuterol | Cetirizine |
| **Leukotriene modifiers** | Levocetirizine |
| Montelukast | Desloratadine |
| Zafirlukast | Diphenhydramine |
| Zileuton | Doxylamine |
| **ICS** | Hydroxyzine |
| Beclomethasone | Chlorpheniramine |
| Budesonide | Dexchlorpheniramine |
| Ciclesonide | Ketotifen |
| Flunisolide | **Leukotriene modifiers** |
| Fluticasone propionate | Montelukast |
| Fluticasone furoate | **Intranasal corticosteroids** |
| Mometasone | Ciclesonide |
| **ICS + LABA** | Fluticasone furoate |
| Budesonide-formoterol | Fluticasone propionate |
| Fluticasone-salmeterol | Mometasone |
| Fluticasone-vilanterol | Beclomethasone |
| Mometasone-formoterol | Budesonide |
| **Asthma Biologics** | Flunisolide |
| Omalizumab | Triamcinolone |
| Mepolizumab | **Intranasal antihistamines** |
| Benralizumab | Azelastine |
| Dupilumab | Olopatadine |
| **OCS** | |
| Prednisone | |
| **Xanthine** | |
| Theophylline | |

*Abbreviations*: ICS, inhaled corticosteroids; ICS+LABA, inhaled corticosteroids + long-acting beta agonist; OCS, oral corticosteroid; SABA, short-acting beta agonist.

**eTable 3. ICD-10-CM codes and scoring system for Charlson Comorbidity Index**

| **Charlson Comorbidity Index**[3] | |
|---|---|
| Myocardial infarction | I21.x, I22.x, I25.2 |
| Congestive heart failure | I11.0, I13.0, I13.2, I25.5, I42.0, I42.5–I42.9, I43.x, I50.x, P29.0 |
| Peripheral vascular disease | I70.x, I71.x, I73.1, I73.8, I73.9, I77.1, I79.0, I79.1, I79.8, K55.1, K55.8, K55.9, Z95.8, Z95.9 |
| Cerebrovascular disease | G45.x, G46.x, H34.0x, H34.1x, H34.2x, I60.x-I68.x |
| Dementia | F01.x-F03.x, F04, F05, F06.1, F06.8, G13.2, G13.8 G30.x, G31.1, G31.2, G91.4, G94, R41.81, R54 |
| Chronic pulmonary disease | J40.x-J47.x, J60.x-J67x, J68.4, J70.1, J70.3 |
| Rheumatic disease | M05.x, M06.x, M31.5, M32.x-M34.x, M35.1, M35.3, M36.0 |
| Peptic ulcer disease | K25.x–K28.x |
| Mild liver disease | B18.x, K70.0-K70.3, K70.9, K71.3–K71.5, K71.7, K73.x, K74.x, K76.0, K76.2, K76.3, K76.4, K76.8, K76.9, Z94.4 |
| Diabetes without chronic complication | Main codes : E08, E09, E10, E11, E13<br>Subcodes : E**.0x, E**.1x, E**.6x, E**.8x, E**.9x |
| Mild to moderate renal disease | I12.9, I13.0, I13.10, N03.x, N05.x, N18.1-4, N18,9, Z94.0 |
| Diabetes with chronic complications | Main codes : E08, E09, E10, E11, E13<br>Subcodes : E**.2, E**.3 E**.4, E**.5 |
| Hemiplegia or paraplegia | G04.1, G11.4, G80.9, G80.1, G80.2, G81.x, G82.x, G83.x |
| Any malignancy | C0x.x, C1x.x, C2x.x, C30.x, C31.x, C32.x, C33.x, C34.x, C37.x, C38.x, C39.x, C40.x, C41.x, C43.x, C45.x, C46.x, C47.x, C48.x, C49.x, C50, C51-58.x, C60-63.x, C76.x, C80.1, C81.x, C82.x, C83.x, C84.x, C85.x, C88.x, C9x.x |
| Moderate to severe liver disease | I85.0x, I86.4, K70.4x, K71.1x, K72.1x, K72.9x, K76.5, K76.6, K67.7 |
| Severe renal disease | I12.0, I13.11, I13.2, N18.5, N18.6, N19.x, N25.0, Z49.x, Z99.2 |
| HIV infection, no AIDS | B20.x |
| Metastatic solid tumor | C77.x-C79.x, C80.0, C80.2 |
| AIDS | B37.x, C53.x, B38.x, B45.x, A07.2, B25.x, G93.4x, B00, B39.x, A07.3, C46.x, C81-C96, A31.x, A15-A19, B59, Z87.01, A81.2, A02.1, B58.x, R64 |
| Charlson comorbidity index[3] | *1 point each:*<br>Myocardial Infarction<br>Congestive heart failure<br>Peripheral vascular disease<br>Cerebrovascular disease<br>Dementia<br>Chronic pulmonary disease<br>Rheumatic disease<br>Peptic ulcer disease<br>Mild liver disease<br>Diabetes without chronic complication<br>Mild to moderate renal disease<br>*2 points each:*<br>Diabetes with chronic complications<br>Hemiplegia or paraplegia<br>Any malignancy<br>*3 points each:*<br>Moderate to severe liver disease<br>Severe renal disease |

| Charlson Comorbidity Index[3] | |
|---|---|
| | HIV infection, no AIDS<br>*6 points each:*<br>Metastatic solid tumor<br>AIDS |

*Abbreviations*: HIV, human immunodeficiency viruses; AIDS, Acquired immunodeficiency syndrome.

**eTable 4. Clinical outcomes of COVID-19 patients with asthma and without asthma: Sensitivity analysis excluding current smokers**

| Outcomes | Asthma (n=532) | No Asthma[a] (n=2542) |
|---|---|---|
| **Hospitalization:** | | |
| No. of events | 114 (21) | 466 (18) |
| Person days of follow up (Mean, SD) | 58 (34) | 61 (33) |
| Rate of event per 1000 person days (95%CI) | 3.7 (3.1, 4.7) | 3.0 (2.8, 3.3) |
| Matched unadjusted (95%CI)[a] | 1.15 (0.95, 1.39) | 1.00 (ref) |
| Partially adjusted hazard ratio (95%CI)[b] | 1.08 (0.89, 1.32) | 1.00 (ref) |
| Fully adjusted hazard ratio (95%CI)[c] | 1.01 (0.82 1,26) | 1.00 (ref) |
| **Mechanical Ventilation:** | | |
| No. of events | 14 (3) | 107 (4) |
| Person days of follow up (Mean, SD) | 71 (23) | 71 (24) |
| Rate of event per 1000 person days (95%CI) | 0.4 (0.2, 0.6) | 0.6 (0.5, 0.7) |
| Matched unadjusted (95%CI)[a] | 0.62 (0.36, 1.08) | 1.00 (ref) |
| Partially adjusted hazard ratio (95%CI)[b] | 0.61 (0.34, 1.08) | 1.00 (ref) |
| Fully adjusted hazard ratio (95%CI)[c] | 0.67 (0.35, 1.26) | 1.00 (ref) |
| **Death:** | | |
| No. of events | 7 (1) | 68 (3) |
| Person days of follow up (Mean, SD) | 73 (21) | 73 (21) |
| Rate of event per 1000 person days (95%CI) | 0.2 (0.0, 0.3) | 0.4 (0.3, 0.5) |
| Matched unadjusted (95%CI)[a] | 0.48 (0.22, 1.04) | 1.00 (ref) |
| Partially adjusted hazard ratio (95%CI)[b] | 0.52 (0.23, 1.17) | 1.00 (ref) |
| Fully adjusted hazard ratio (95%CI)[c] | **0.31 (0.12, 0.83)** | 1.00 (ref) |

Abbreviations: COVID-19, coronavirus disease 2019; SD, standard deviation; CI, confidence interval.
Text in boldface indicates statistical significance, *p* <0.05.).
[a]Age, sex, and date of SARS-CoV-2 diagnosis matched comparators.
[b] Matched on age, sex, and date of SARS-CoV-2 test date and adjusted for age, sex, race, ethnicity, payor, and smoking status.
[c] Matched on age, sex, and date of SARS-CoV-2 test date and adjusted for age, sex, race, ethnicity, payor, smoking status, body mass index, and Charlson comorbidity index.

**eTable 5. Characteristics of COVID-19 patients with allergic asthma and non-allergic asthma**

| Characteristic | Allergic Asthma (n=260) | No Asthma (n= 1252) | *P*-Value[a] | Non-Allergic Asthma (n=302) | No Asthma (n = 1434) | *P*-Value[b] |
|---|---|---|---|---|---|---|
| Age (mean, SD, years) | 52 (16) | 51 (16) | - | 51 (17) | 50 (17) | - |
| 26-35 | 35 (13) | 170 (14) | | 44 (15) | 218 (15) | |
| 36-45 | 59 (23) | 290 (20) | | 56 (19) | 274 (19) | |
| 46-55 | 51 (20) | 251 (20) | | 76 (25) | 357 (25) | |
| 56-65 | 55 (21) | 260 (21) | | 71 (24) | 338 (24) | |
| 66-75 | 42 (16) | 196 (16) | | 27 (9) | 126 (9) | |
| 76-85 | 15 (6) | 70 (6) | | 17 (6) | 69 (5) | |
| 86-95 | 2 (1) | 10 (1) | | 10 (3) | 47 (3) | |
| >95 | 1 (< 1) | 5 (< 1) | | 1 (< 1) | 5 (< 1) | |
| Female | 210 (81) | 1009 (81) | - | 194 (64) | 924 (64) | - |
| Race | | | 0.32 | | | **0.001** |
| White | 126 (48) | 580 (46) | | 177 (59) | 667 (47) | |
| Black | 48 (18) | 225 (18) | | 47 (16) | 250 (17) | |
| Asian | 4 (2) | 48 (4) | | 6 (2) | 52 (4) | |
| Other | 82 (32) | 399 (32) | | 72 (24) | 465 (32) | |
| Ethnicity | | | **0.01** | | | **< 0.001** |
| Hispanic | 92 (35) | 378 (30) | | 84 (28) | 478 (33) | |
| Non-Hispanic | 158 (61) | 763 (61) | | 210 (70) | 832 (58) | |
| Unknown | 10 (4) | 111 (9) | | 8 (3) | 124 (9) | |
| Payor | | | **0.003** | | | **0.002** |
| Private | 156 (60) | 732 (58) | | 187 (62) | 852 (59) | |
| Public | 102 (39) | 449 (36) | | 111 (37) | 488 (34) | |
| Other | 2 (1) | 71 (6) | | 4 (1) | 94 (7) | |
| Body mass index (mean, SD, $kg/m^2$) | 32.9 (7.4) | 29.8 (6.5) | **< 0.001** | 30.8 (7.0) | 29.7 (6.3) | **0.01** |
| <18.5 | 4 (2) | 306 (24) | **< 0.001** | 4 (1) | 375 (26) | **< 0.001** |
| 18.5-24.9 | 26 (10) | 217 (17) | | 51 (17) | 242 (17) | |
| 25-29.9 | 58 (22) | 314 (25) | | 99 (33) | 345 (24) | |
| 30-34.9 | 84 (32) | 229 (18) | | 76 (25) | 274 (19) | |
| 35-39.9 | 50 (19) | 116 (9) | | 46 (15) | 133 (9) | |
| ≥40 | 38 (15) | 70 (6) | | 26 (9) | 65 (5) | |
| Smoking status | | | **< 0.001** | | | **< 0.001** |
| Never | 184 (71) | 763 (61) | | 201 (67) | 858 (60) | |
| Former | 64 (25) | 205 (16) | | 77 (26) | 227 (16) | |

| Characteristic | Allergic Asthma (n=260) | No Asthma (n= 1252) | *P*-Value[a] | Non-Allergic Asthma (n=302) | No Asthma (n = 1434) | *P*-Value[b] |
|---|---|---|---|---|---|---|
| Current | 10 (4) | 47 (4) | | 20 (7) | 54 (4) | |
| Unknown | 2 (1) | 237 (19) | | 4 (1) | 295 (21) | |
| Diabetes | 48 (18) | 106 (8) | **< 0.001** | 51 (17) | 116 (8) | **< 0.001** |
| Hypertension | 18 (7) | 34 (3) | **0.001** | 18 (6) | 37 (3) | **0.002** |
| Congestive heart failure | 7 (3) | 13 (1) | 0.06 | 6 (2) | 17 (1) | 0.27 |
| Severe renal disease | 2 (1) | 10 (1) | 1.00 | 5 (2) | 13 (1) | 0.22 |
| Sickle cell disease | 2 (1) | 1 (< 1) | 0.08 | 1 (< 1) | 4 (< 1) | 1.00 |
| Organ transplantation | 0 (0) | 0 (0) | - | 0 (0) | 0 (0) | - |
| Charlson comorbidity index[c] | 2.1 (2.2) | 1.1 (2.3) | **< 0.001** | 2.1 (2.8) | 0.9 (1.9) | **< 0.001** |
| Asthma medications | | | | | | |
| SABA | 232 (89) | 116 (9) | **< 0.001** | 253 (84) | 132 (9) | **< 0.001** |
| Montelukast | 77 (30) | 5 (< 1) | **< 0.001** | 0 (0) | 3 (< 1) | 1.00 |
| ICS | 173 (67) | 35 (3) | **< 0.001** | 150 (50) | 24 (2) | **< 0.001** |
| ICS + LABA | 99 (38) | 10 (< 1) | **< 0.001** | 73 (24) | 9 (< 1) | **< 0.001** |
| Biologic[d] | 7 (3) | 2 (< 1) | **< 0.001** | 1 (< 1) | 0 (0) | 0.17 |
| OCS | 18 (7) | 13 (1) | **< 0.001** | 22 (7) | 9 (< 1) | **< 0.001** |
| Theophylline | 0 (0) | 1 (< 1) | 1.00 | 2 (< 1) | 0 (0) | **0.03** |
| None | 0 (0) | 1109 (89) | **< 0.001** | 0 (0) | 1279 (89) | **< 0.001** |

*Abbreviations*: COVID-19, coronavirus disease 2019; SD, standard deviation; SABA, short acting beta agonists; ICS, inhaled corticosteroid; ICS +LABA, inhaled corticosteroid + long-acting beta agonist; LAMA, long acting muscarinic antagonist; OCS, oral corticosteroid.
Data are represented by mean ± SD or number (%) unless otherwise indicated. Text in boldface indicates statistical significance, p <0.05.
[a] *P*-value comparing allergic asthma to no asthma.
[b] *P*-value comparing non-allergic asthma to no asthma.
[c] See **eTable 3** for ICD-10-CM codes and scoring system.
[d] Biologic therapy includes anti-IgE, anti-IL5/IL5R, anti-IL4R.

### eTable 6. Characteristics of COVID-19 patients with severe and non-severe asthma

| Characteristic | Severe Asthma (n=44) | No Asthma (n= 210) | P-value[a] | Non-Severe (n=518) | No Asthma (n = 2476) | P-value[b] |
|---|---|---|---|---|---|---|
| Age (mean, SD, years) | 54 (17) | 53 (17) | - | 51 (17) | 51 (17) | - |
| 26-35 | 4 (9) | 18 (9) | | 75 (14) | 370 (15) | |
| 36-45 | 11 (25) | 54 (26) | | 104 (20) | 510 (21) | |
| 46-55 | 12 (27) | 58 (28) | | 115 (22) | 550 (22) | |
| 56-65 | 5 (11) | 25 (12) | | 121 (23) | 573 (23) | |
| 66-75 | 6 (14) | 30 (14) | | 63 (12) | 292 (12) | |
| 76-85 | 4 (9) | 15 (7) | | 28 (5) | 124 (5) | |
| 86-95 | 2 (5) | 10 (5) | | 10 (2) | 47 (2) | |
| >95 | - | - | | 2 (< 1) | 10 (< 1) | |
| Female | 33 (75) | 155 (74) | - | 371 (72) | 1778 (72) | - |
| Race | | | **0.04** | | | **0.003** |
| White | 29 (66) | 105 (50) | | 274 (53) | 1142 (46) | |
| Black | 2 (5) | 38 (18) | | 93 (18) | 437 (18) | |
| Asian | 2 (5) | 3 (1) | | 8 (2) | 97 (4) | |
| Other | 11 (25) | 64 (2303) | | 143 (28) | 800 (32) | |
| Ethnicity | | | 0.56 | | | **< 0.001** |
| Hispanic | 12 (27) | 57 (27) | | 164 (32) | 799 (32) | |
| Non-hispanic | 30 (68) | 133 (63) | | 338 (65) | 1462 (59) | |
| Unknown | 2 (5) | 20 (10) | | 16 (3) | 215 (9) | |
| Payor | | | 0.41 | | | **< 0.001** |
| Private | 27 (61) | 127 (60) | | 316 (61) | 1457 (59) | |
| Public | 17 (39) | 75 (36) | | 196 (38) | 862 (35) | |
| Other | 0 (0) | 8 (4) | | 6 (1) | 157 (6) | |
| Body mass index (mean, SD, kg/m$^2$) | 30.9 (6.9) | 29.6 (6.7) | 0.26 | 31.9 (7.3) | 29.8 (6.4) | **< 0.001** |
| <18.5 | 1 (2) | 43 (20) | **0.04** | 7 (1) | 638 (26) | **< 0.001** |
| 18.5-24.9 | 7 (16) | 38 (18) | | 70 (14) | 421 (17) | |
| 25-29.9 | 14 (32) | 59 (28) | | 143 (28) | 600 (24) | |
| 30-34.9 | 10 (23) | 41 (20) | | 150 (29) | 462 (19) | |
| 35-39.9 | 7 (16) | 18 (9) | | 89 (17) | 231 (9) | |
| >40 | 5 (11) | 11 (5) | | 59 (11) | 124 (5) | |
| Smoking status | | | **0.03** | | | **< 0.001** |
| Never | 33 (75) | 124 (59) | | 352 (68) | 1497 (60) | |
| Former | 10 (23) | 45 (21) | | 131 (25) | 387 (16) | |
| Current | 1 (2) | 8 (4) | | 29 (6) | 93 (4) | |
| Unknown | 0 (0) | 33 (16) | | 6 (1) | 499 (20) | |
| Diabetes | 10 (23) | 19 (9) | **0.01** | 89 (17) | 203 (8) | **< 0.001** |
| Hypertension | 8 (128 | 4 (2) | **< 0.001** | 28 (5) | 67 (3) | **0.002** |
| Congestive heart failure | 3 (7) | 3 (1) | 0.07 | 10 (2) | 27 (1) | 0.12 |
| Severe renal disease | 3 (7) | 1 (< 1) | **0.02** | 4 (< 1) | 22 (< 1) | 1.00 |
| Sickle cell disease | 0 (0) | 1 (< 1) | 1.00 | 3 (< 1) | 4 (< 1) | 0.10 |
| Organ transplantation | 0 (0) | 0 (0) | - | 0 (0) | 0 (0) | - |

| Characteristic | Severe Asthma (n=44) | No Asthma (n= 210) | P-value[a] | Non-Severe (n=518) | No Asthma (n = 2476) | P-value[b] |
|---|---|---|---|---|---|---|
| Charlson comorbidity index[c] | 3.5 (4.0) | 1.0 (1.8) | **< 0.001** | 2.0 (2.3) | 1.0 (2.1) | **< 0.001** |
| Asthma medications | | | | | | |
| SABA | 33 (75) | 18 (9) | **< 0.001** | 452 (87) | 230 (9) | **< 0.001** |
| Montelukast | 10 (23) | 0 (0) | **< 0.001** | 67 (13) | 8 (< 1) | **< 0.001** |
| ICS | 30 (68) | 4 (2) | **< 0.001** | 239 (57) | 55 (2) | **< 0.001** |
| ICS + LABA | 21 (48) | 2 (0) | **< 0.001** | 151 (29) | 17 (< 1) | **< 0.001** |
| Biologic[d] | 8 (18) | 0 (< 1) | **< 0.001** | 0 (0) | 2 (< 1) | 1.00 |
| OCS | 40 (91) | 3 (1) | **< 0.001** | 0 (0) | 19 (< 1) | 0.06 |
| Theophylline | 1 (2) | 0 (0) | 0.17 | 1 (< 1) | 1 (< 1) | 0.32 |
| None | 0 (0) | 189 (90) | **< 0.001** | 0 (0) | 2199 (89) | **< 0.001** |

Abbreviations: COVID-19, coronavirus disease 2019; SD, standard deviation; SABA, short acting beta agonists; ICS, inhaled corticosteroid; ICS +LABA, inhaled corticosteroid + long-acting beta agonist; LAMA, long acting muscarinic antagonist; OCS, oral corticosteroid.

Data are represented by mean ± SD or number (percentage) unless otherwise indicated. Text in boldface indicates statistical significance, $p < 0.05$.

[a] P-value comparing severe asthma to no asthma.

[b] P-value comparing non-severe asthma to no asthma.

[c] See **eTable 3** for ICD-10-CM codes and scoring system.

[d] Biologic therapy includes anti-IgE, anti-IL5/IL5R, anti-IL.
